# Implementation of culturally responsive communication for racial, ethnic, sexual, and gender minoritized patients when screened for COVID-19 vaccinations: A scoping review

**DOI:** 10.1101/2025.03.30.25324915

**Authors:** Nikhil Kalita, Patrick G Corr, Maranda C Ward, Julia Xavier, Paige L McDonald

## Abstract

**Introduction:** The COVID-19 pandemic exacerbated long-standing healthcare disparities, disproportionately affecting racial, ethnic, sexual, and gender minoritized populations. Structural inequities fuel medical mistrust and hinder equitable vaccine access. Culturally responsive communication (CRC) is a critical strategy in primary care that has the potential to improve patient-provider interactions and vaccine acceptance.

**Objectives:** This scoping review examines how CRC is conceptualized and implemented in clinical interactions related to COVID-19 vaccination and booster screening for minoritized populations. It assesses the scope of CRC research, the clarity of definitions, the extent of implementation, and its clinical applicability.

**Methods:** Following Arksey and O’Malley’s framework and PRISMA-ScR guidelines, we conducted a literature search across four databases, analyzing studies published between November 2019 and 2022. Extracted data included CRC definitions, communication strategies, and interventions from 22 eligible studies.

**Results:** Research on CRC in the context of COVID-19 vaccination is limited and inconsistent. Most studies focused on Black and Hispanic populations, with a critical gap in research addressing sexual and gender minorities. CRC terminology was often interchangeable with concepts like cultural competence, leading to definitional inconsistencies. Because public health messaging was a primary focus, direct clinical applications of CRC were underexplored.

**Discussion:** Our findings highlight an urgent need for a standardized CRC framework to enhance healthcare equity. The absence of a clear, universally accepted definition hinders CRC’s practical application and measurability in clinical settings. Future research should refine CRC conceptualization, establish measurable interventions, and expand inclusivity to sexual and gender minorities to foster more equitable healthcare practices.

## Introduction

The coronavirus disease 2019 (COVID-19) pandemic shed light on the impact of racism, heterosexism, and transphobia within healthcare settings.[1–2] COVID-19 disproportionately impacts racial, ethnic, sexual, and gender minoritized communities due to systemic barriers and structural inequities.[1–2] Racial and ethnic minorities, for example, face higher COVID-19 infection rates and mortality compared to white populations.[3] Structural disadvantages—such as limited healthcare access, racism, and environmental stressors—are known contributors to this disparity.[4] Similarly, sexual and gender minority groups, although underrepresented in COVID-19 data, report heightened mental health challenges, economic insecurity, and minority stress, further increasing their vulnerability.[2, 5–10]

Disparities in COVID-19 vaccine access and acceptance contributed to these inequities.[11] Initial vaccine uptake was lower among racial and ethnic minorities compared to white populations, and sexual and gender minorities experienced unique barriers linked to historical medical trauma.[2–3, 12] This mistrust stems from historical abuses, such as nonconsensual medical experimentation on Black Americans, stigmatization in the human immunodeficiency virus (HIV) crisis, and current healthcare practices.[1,13–15] These factors highlight the urgent need for culturally responsive communication (CRC) from primary care providers (PCPs), especially during public health crises like COVID-19.[2,16–17]

Communication disparities between PCPs and minoritized patients persist, often perpetuating existing medical mistrust among minoritized populations.[1,18–22] CRC plays a critical role in patient trust and vaccine acceptance, positioning PCPs as trusted sources of information.[17,23–25] Given the importance of CRC for addressing health disparities, it is essential to understand its current implementation in PCP interactions with minoritized patients, especially regarding COVID-19 vaccination screening.[16–17]

Both COVID-19 and HIV are highly stigmatized, preventable, and communicable viruses that disproportionately affect health outcomes for minoritized groups.[15,26] Utilizing CRC in screenings for COVID-19 and HIV in the same primary care setting could help alleviate the negative impact of the various structural and interpersonal barriers to care.[2,27] To facilitate simultaneous culturally responsive screening for COVID-19 and HIV, our study team developed training modules to enhance PCP’s capacity in CRC.[28–30] We conducted two distinct scoping reviews to guide the design of this training, along with corresponding social marketing campaigns and policy recommendations.[31–33] The protocol for this scoping review has been recently published and below we present the culmination of this research.[33]

## Objectives

The primary research question guiding this scoping review was, “How is CRC occurring between patient and practitioner related to COVID-19 vaccination and booster screening for racial, ethnic, sexual, and gender minoritized patients?” To answer this question, we analyzed the literature with the four sub-questions below:

1. How is culturally responsive communication being researched in clinical interactions regarding COVID vaccinations among minoritized groups?
2. At what level is culturally responsive communication occurring (public health communication, interpersonal patient-PCP communication, etc.)?
3. Is culturally responsive communication being adequately defined in each of these studies, and if so, how is it being defined or at least recommended?
4. Is culturally responsive communication measurable in clinical settings?

## Methods

As detailed in our published protocol, this study followed the framework for conducting scoping reviews as established by Arksey and O’Malley and the PRISMA-ScR guidelines from JBI.[34–35] The five-stage process guiding this study is detailed below.

## Stage 1: Identifying the Research Question

Before commencing the scoping review, the research team formulated the initial research question after consulting with primary care physicians (PCPs). These consultations emphasized the importance of focusing on CRC concerning COVID-19 vaccinations rather than general COVID-19 prevention and screening. Given the ongoing challenges with COVID-19 vaccine acceptance and the disproportionate impact on minoritized populations, we crafted the research question above.

## Stage 2: Identifying Relevant Studies

The search for relevant studies was conducted across four databases: MEDLINE (Pubmed), Scopus, CENTRAL (Cochrane Central Registry of Controlled Trials), and CINAHL (Complete). Collaborating with research team members and an experienced librarian, an inclusive search strategy was developed, encompassing various terms related to the research question and key concepts.

The search strategy involved four main categories of terms: minoritized patient groups in the U.S., related terms of CRC, COVID-19, and vaccination. To ensure inclusivity, terms encompassing different terminology and synonyms for all categories were considered. These terms are comprehensively detailed in the published protocol.[33]

The search yielded varying results across databases: 284 results from MEDLINE, 545 from SCOPUS, 61 reviews and 114 prospective clinical trials from CENTRAL, and 127 results from CINAHL. Despite encountering formatting and character technicalities, adjustments were made to ensure consistency and accuracy across searches.

## Stage 3: Study/Article Selection

The scoping review employs Covidence literature review software for two levels of screening. Initially, titles and abstracts were screened by four independent investigators based on predetermined inclusion criteria (NK, PC, PM, JX). Studies meeting these criteria were included in full-text screening, where inclusion decisions are made by two reviewers. In cases of disagreement, a third reviewer provided the final decision. Both stages of screening adhered to specific eligibility criteria related to study location, focus on racial, ethnic, sexual, and gender minoritized groups, inclusion of COVID-19 vaccine-related content, relevance to patient and PCP attitudes or behaviors, publication after November 2019, and exclusion of unpublished research or protocols.

### Inclusion Criteria

The search was limited to articles with the following characteristics:

● Articles concerned screening services, specifically regarding COVID vaccination screening, among racial, ethnic, sexual, and gender minoritized patient populations,
● Articles published in peer-reviewed research journals,
● Articles published in English,
● Articles addressing the issue of COVID-19 vaccine screening in the context of the United States healthcare systems.[33]

### Exclusion Criteria

Articles were excluded that met the following exclusion criteria:

● Articles for which full-text publications were not available,
● Book chapters and study protocols,
● Articles published in a language other than English,
● Articles published prior to 2019,
● Articles including data solely collected from any country other than the U.S., except for systematic reviews,
● Articles that do not address one or more of our historically marginalized populations of interest or studies that do not discuss COVID-19 vaccination screening.[33]

## Stage 4: Charting the Data

Data extraction was facilitated through Google Sheets to enhance collaboration and flexibility. Nine reviewers utilize an evidence-based format with prompts and questions for extracting information from included studies (AK, MW, HC, NK, PC, PM, OC, MCW, PD). These prompts cover various aspects such as study design, intervention types, study population demographics, methodology overview, results, communication levels addressed, and considerations for racial, ethnic, sexual, and gender minoritized groups. The extracted data was organized systematically in Google Sheets for further analysis. To ensure rigor, three primary reviewers conducted an additional review of all studies to assess substantial mention of direct CRC between PCPs and patients (NK, PM, PC). If CRC data was found to be present, the studies were included for further analysis based on predetermined criteria. The results of this process are summarized below and the full dataset is available upon request.

## Stage 5: Collating, Summarizing, and Reporting the Results

The collated data was organized thematically and assessed for relevance to the research question, focusing on CRC related to COVID-19 vaccination for minoritized patients. Themes emerged through inductive coding and were analyzed through the lenses of Critical Race Theory,[36] Queer Theory,[37] and the Socio-Ecological Framework (SEM).[38] The review prioritizes identifying the breadth of available literature over quality assessment typically done in systematic reviews. Findings were synthesized and reported following PRISMA-ScR guidelines, including a PRISMA flowchart (Figure 1) detailing the study selection process. The results will be disseminated to inform future research and interventions regarding CRC for COVID-19 vaccination screening in primary care settings.

**Figure 1.**
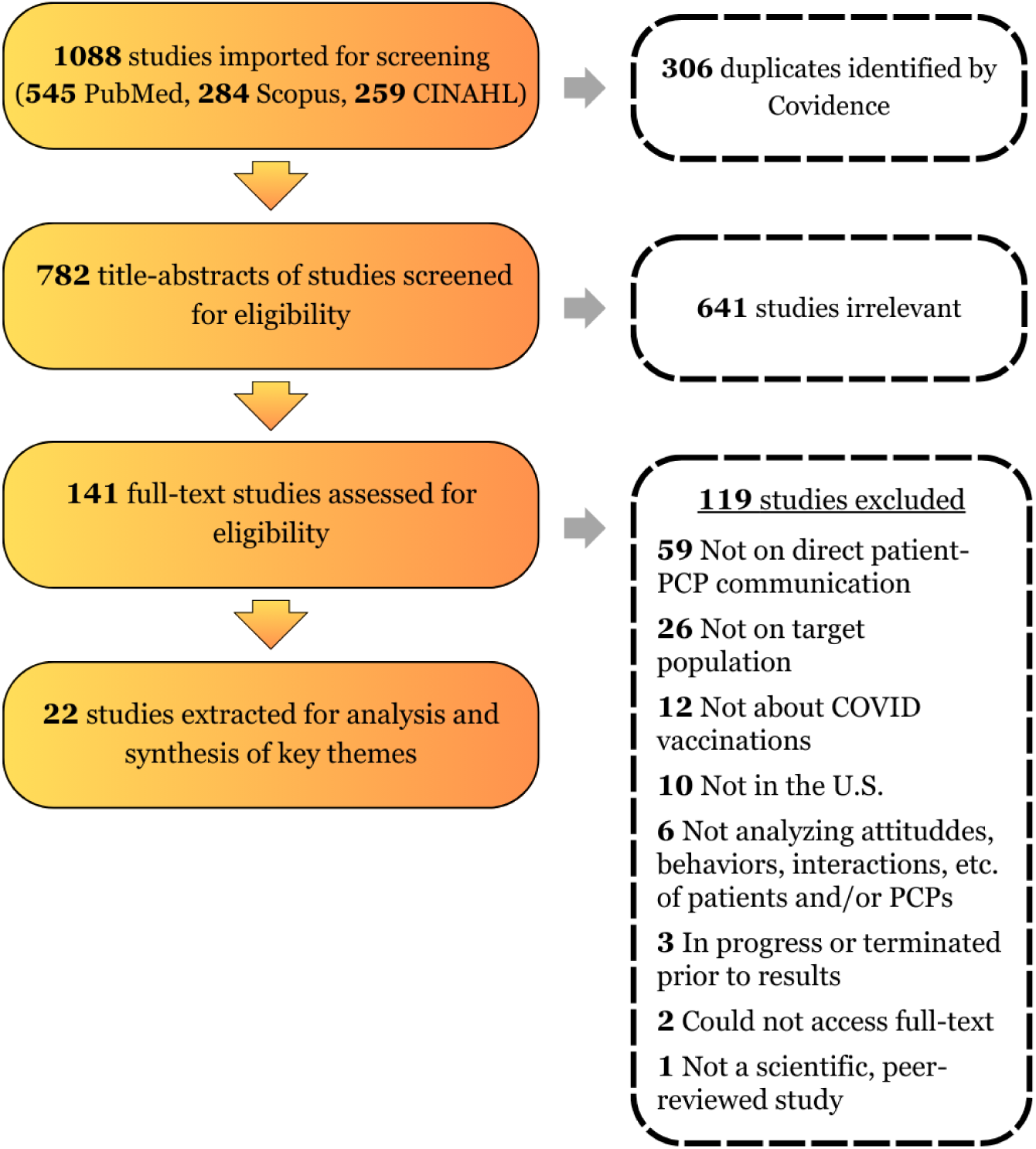
PRISMA Diagram

## Results

Findings are divided into two distinct sections. First we present an overview of study characteristics. Next, we present results related to our specific primary and sub-research questions.

## Study Characteristics

Prior to analyzing the salient results from each study evaluated for this scoping review, we collated data on the study characteristics. This data provides an overview of the year of publication, the type of research conducted, and the populations studied across the 22 articles included in this review. This data is presented in Tables 1-4.

**Table 1:** Study counts by year of study or intervention.

| **Year of Publication** | **Count** | **Citation Number** |
| --- | --- | --- |
| 2021 | 6 | 39, 40, 56, 57, 58, 59 |
| 2022 | 16 | 41, 42, 43, 44, 45, 46, 47, 48, 49, 50, 51, 52, 53, 54, 55, 60 |

**Table 2:** Study counts by article methodology types.

| **Article Type** | **Count** | **Citation Number** |
| --- | --- | --- |
| Empirical Research | 13 | 39, 42, 44, 45, 46, 48, 49, 50, 52, 53, 55, 59, 60 |
| Literature Review | 4 | 40, 41, 43, 56 |
| Commentary | 3 | 54, 57, 58 |
| Conceptual Articles | 2 | 47, 51 |

**Table 3:** Study counts by populations of focus.

| **Population** | **Count** | **Citation Number** |
| --- | --- | --- |
| Black or African American | 9 | 41, 43, 44, 48, 51, 52, 54, 55, 58 |
| Hispanic/Latino American | 2 | 45, 49 |
| Multiple Populations (BIPOC) | 11 | 39, 40, 42, 46, 47, 50, 53, 56, 57, 59, 60 |

**Table 4:** Study counts by level of communication.

| **Level** | **Count** | **Citation Numbers** |
| --- | --- | --- |
| Online Patient-PCP Communication | 2 | 41, 50 |
| In-Person Patient-PCP Communication | 12 | 39, 40, 42, 44, 45, 46, 47, 49, 52, 56, 57, 58 |
| Public health<br>Communication | 8 | 43, 48, 51, 53, 54, 55, 59, 60 |

The majority of articles included in the scoping review were published in 2022. The increase in publication could relate to the continued trend in the COVID pandemic and the evolving public health landscape. We expect that increasing the inclusion criteria to incorporate articles released in 2023 and 2024 would further increase this number.

The articles under review ranged in types. Of the 22 articles, 13 articles involved reports of research findings: seven articles were based on qualitative studies,[45, 48–49, 52, 55, 59–60] and one article was a quantitative survey study.[39] One research study was based on education research.[44] Two studies included participatory action or community-based research.[46, 53] Two studies were reports of public health interventions with components involving clinical interactions.[42, 50] Of the remaining nine articles, four were literature reviews,[40–41, 43, 56] three were opinion pieces or commentaries[54, 57–58] and two were conceptual articles.[47, 51]

This review specifically considered publications related to CRC between clinicians and minoritized populations. Table 3 provides an overview of these characteristics. Of note, eleven of the articles included a diverse sampling of participants from multiple racial backgrounds.[39–40, 42,46–47, 50, 53, 56–57, 59–60] This typically meant that the authors of the study spoke in broad terms about patient populations from racial, ethnic, sexual, or gender minoritized backgrounds without specifically stratifying by specific population.

## Level of Communication

Because of the nature of our question and construction of our search, all articles in the review addressed some aspect of communication among patients and providers. But not all articles addressed interpersonal patient-PCP communication. Even with the rigor of our search strategy, during analysis of resulting articles, we identified articles focused on public health communication from patient to provider rather than direct interpersonal patient-PCP communication. As a consequence, we developed the fourth sub question guiding our analysis regarding at what level the patient-PCP communication occurring. As Table 4 indicates, there were several levels of communication present in our review: online patient-PCP communication, in-person patient-PCP communication, and public health communication, with studies measuring or recommending cultural responsiveness at varying levels. Twelve articles investigated and recommended cultural responsiveness for PCPs who are in face-to-face contact with patients.[39–40, 42, 44, 47, 49, 52, 56–58] In two of the articles, the patient-PCP communication was through indirect, online contact.[41, 50] These studies focused on developing or recommending PCP cultural responsiveness through online communication with patients. In eight articles, cultural responsiveness was measured or recommended based on public health communication campaigns.[43, 48, 51, 53–55, 59–60] These studies had a particular focus on identifying community needs to develop culturally responsive interventions in health care systems and community-based clinics. Interventions demonstrated or recommended in these articles involved some type of interpersonal cultural responsiveness between PCPs and patients such as cultural and language concordance,[43, 45–46, 53, 57–58] PCP postcard communication that addresses cost and immigration status,[50] and culturally tailored social media vaccine outreach.[49]

## CRC Defined

One interesting finding related to the scoping review question is that while all articles included in the review either mentioned culturally responsive communication or a related concept (cultural competence, humility, tailoring, sensitivity, concordance, etc.), not all provided clear and consistent definitions or defined them in the context of a specific community (i.e., by racial, ethnic, sexual, and gender identity). Table 5 indicates the reviewed article’s citation number, the terminology used within articles, the definitions of CRC provided, or if not provided, how CRC was recommended in the article.

**Table 5:**
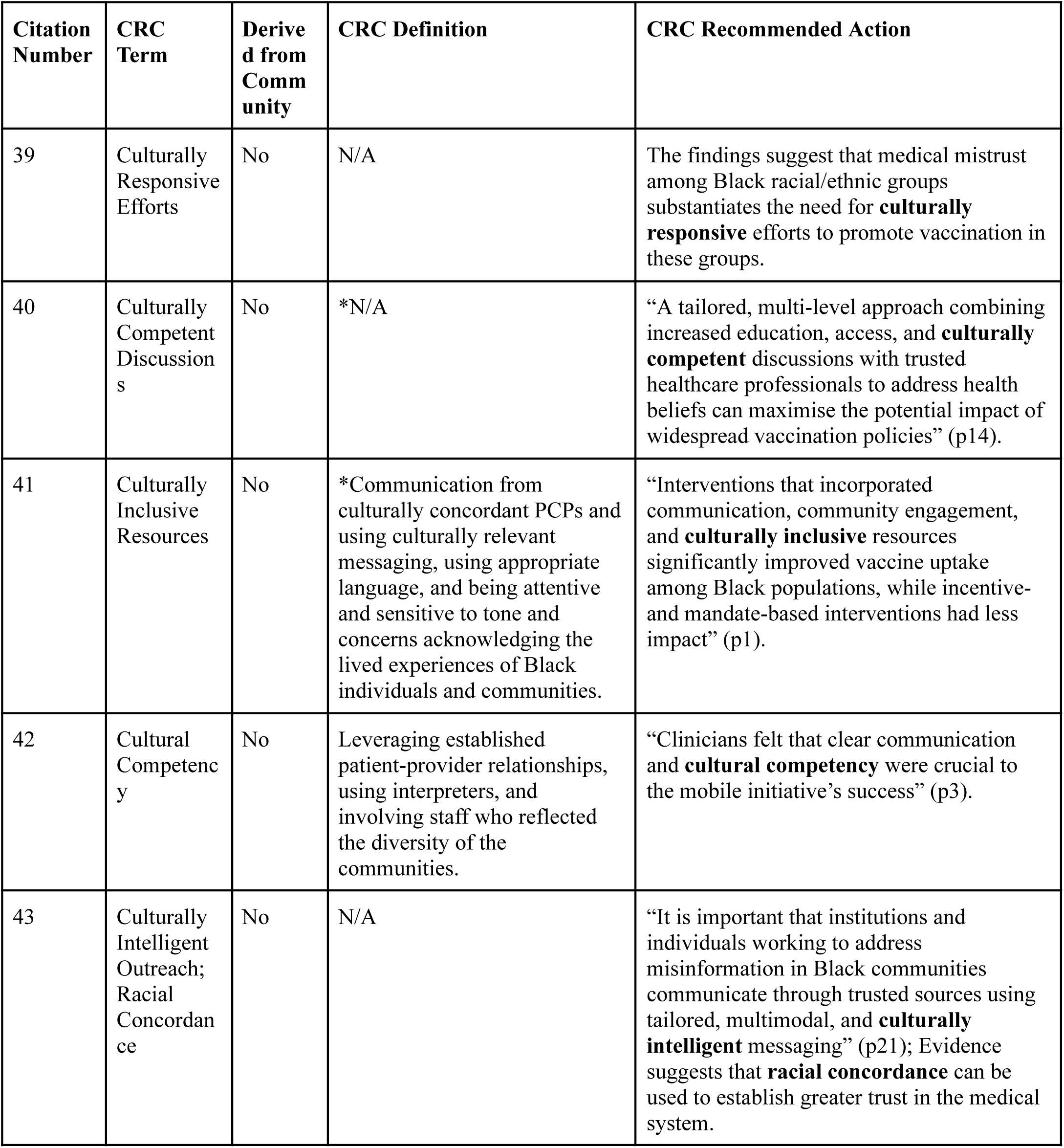
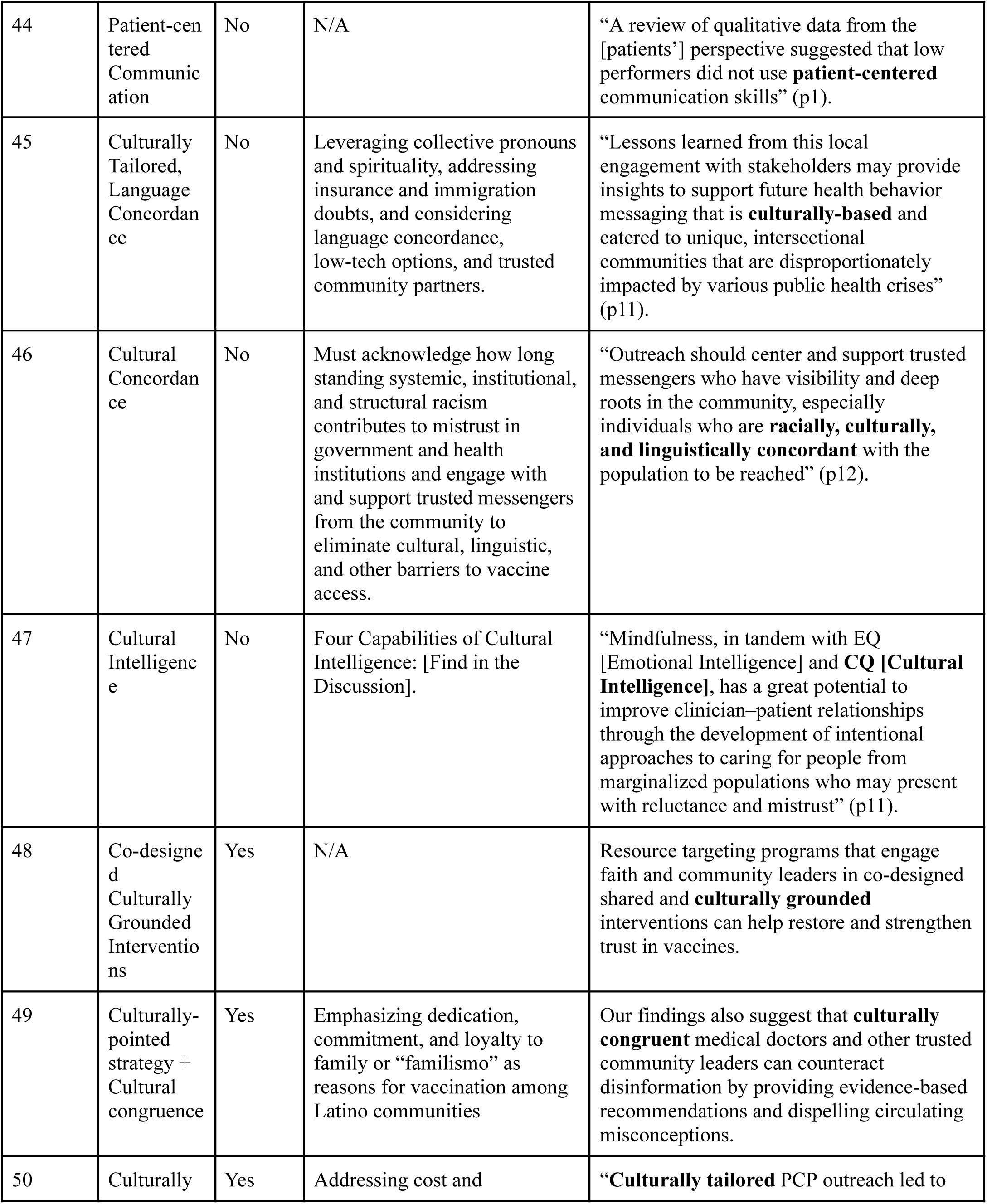
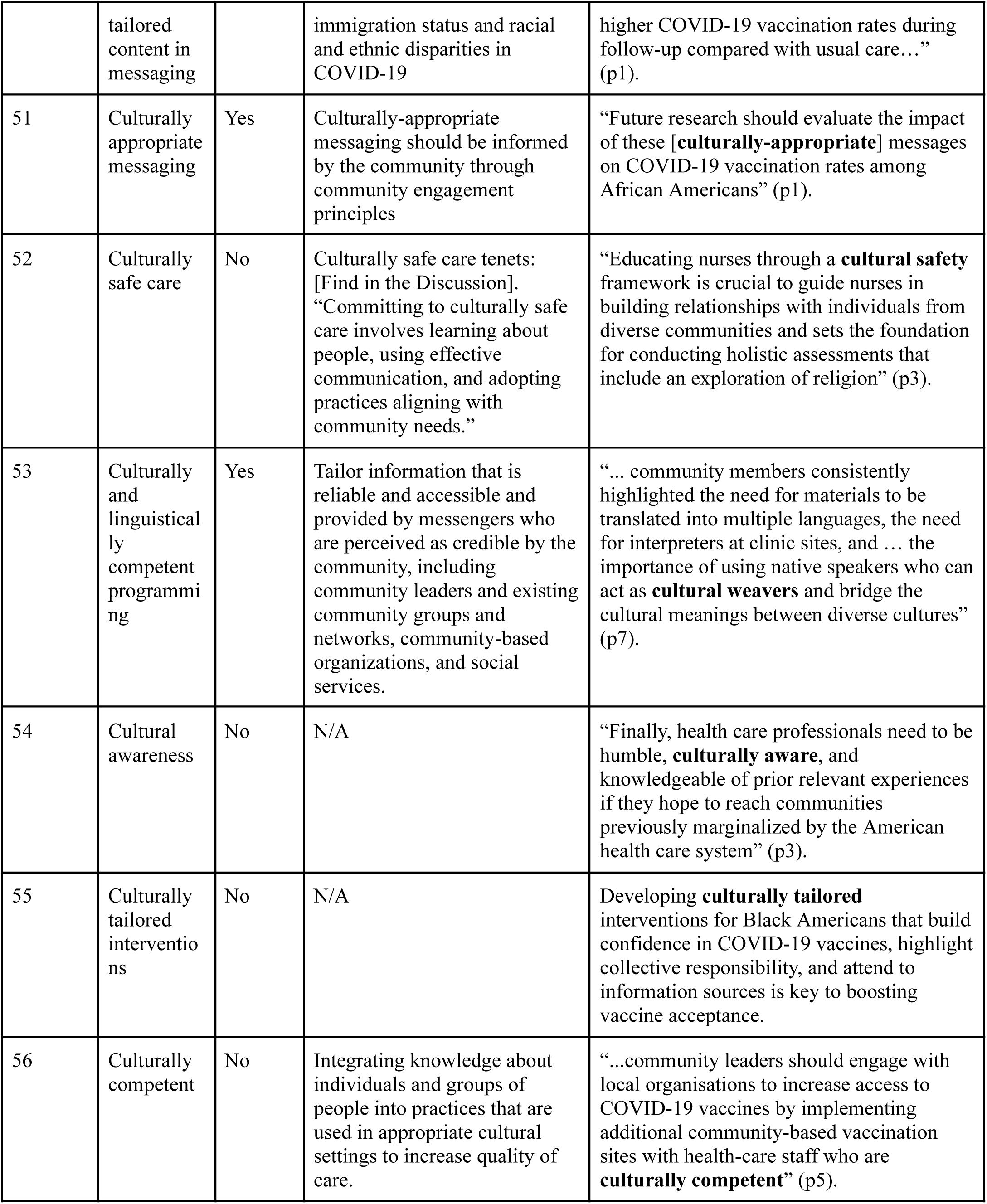
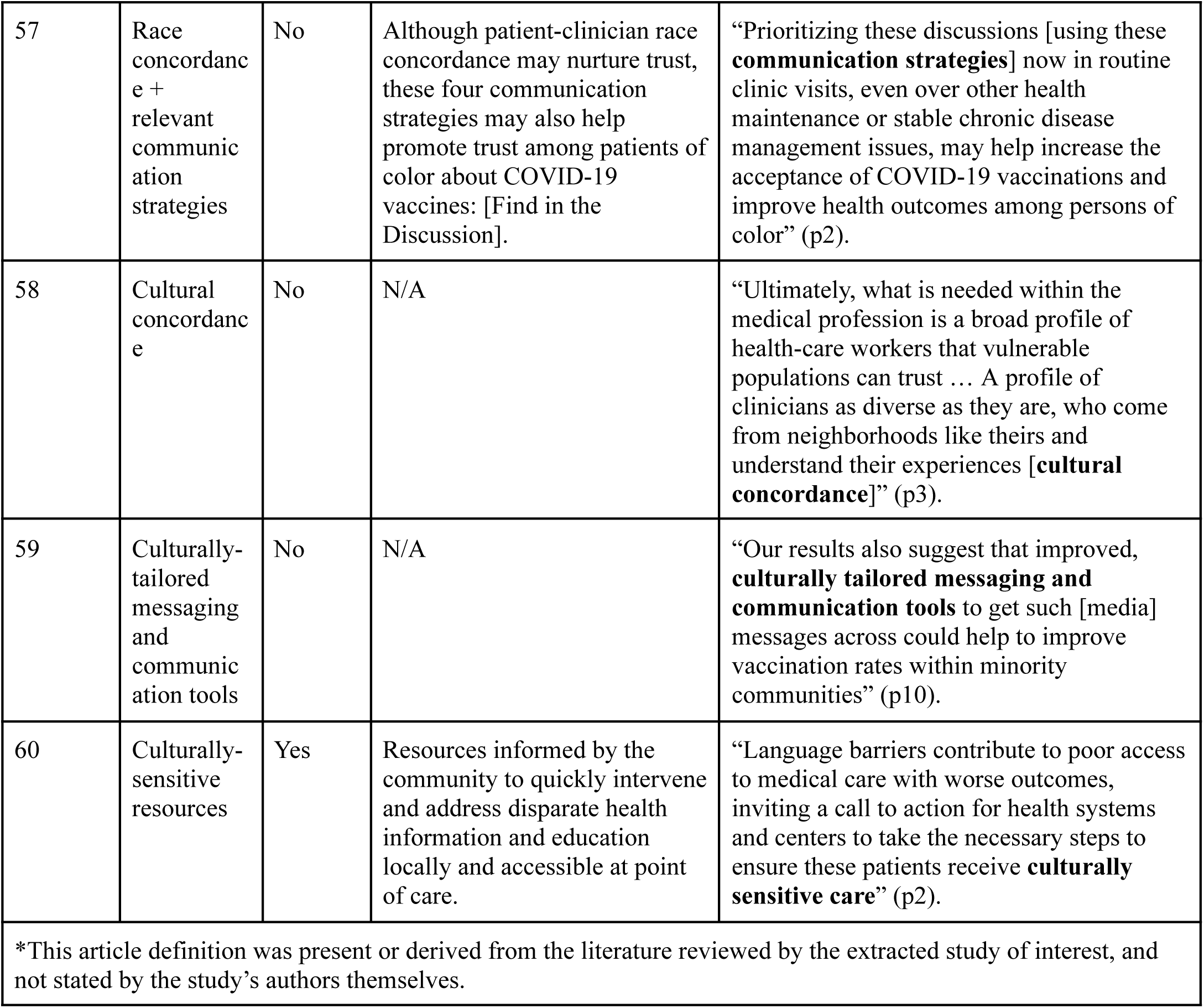
CRC related terms, community derivation, definitions, and recommendations by citation number.

Eight articles provided definitions of related concepts that can be generalized for all communities.[42, 45–47, 52, 56–58] For example, Omer et al. defined the commonly utilized term, cultural competence, as PCPs “who integrate knowledge about individuals and groups of people into practices that are used in appropriate cultural settings to increase the quality of care”.^[56(p1)]^ Some articles defined their CRC-related concept through their description of interventions, like Light et al. who described that “culturally tailored messaging may benefit from leveraging collective pronouns and spirituality, addressing insurance and immigration doubts, and considering language concordance, low-tech options, and trusted community partners”.^[45(p11)]^ One of the standout articles that solely focused on defining a CRC-related concept was by Richard-Eagline & McFarland where they created a framework with four detailed capabilities that comprehensively and intuitively defined their idea of cultural intelligence.[47]

Seven articles defined cultural responsiveness for individual communities based on extracting specific community perspectives.[48–51, 53, 55, 60] These studies often involved community-based qualitative research to determine the specific needs for their community of interest and to address them via recommendations or direct intervention. For example, Stadnick et al. utilized a participatory action research design to create “Community Advisory Boards” that “focused on identifying necessary conditions that must exist to eliminate COVID-19 disparities” for specific Latino, Black, and immigrant and refugee communities in San Diego.[53] These community members repeatedly indicated the need for clinic materials to be translated in multiple languages and the need for interpreters at clinic sites, which led to the development of the necessary condition of linguistically and culturally competent programming. Another study by Majee et al. derived their ideas of “context-based culturally sensitive interventions” from directly interviewing community church participants and leaders who were primarily African American.[48] They found that some of the experiences this African American community had during hospital visits resurrected their feelings of racism, but other interpersonal and institutional experiences have built their confidence in their PCPs. This finding directly highlights the study’s recommendation for a culturally competent clinical environment.

Six articles mentioned or recommended CRC-related concepts, but did not define them.[39–40, 43, 48, 54, 59] For example, at the end of the article by Alrassi et al., authors state that “health care professionals need to be humble, culturally aware, and knowledgeable of prior relevant experiences if they hope to reach communities previously marginalized by the American health care system”.^[54(p3)]^ Here, they implicate the need for PCPs to be culturally aware, but do not define how they are supposed to be culturally aware. This was especially common for the literature review studies where the studies they reviewed may have defined the CRC-related terms, but the literature review itself only recommended them. The remaining three articles did not name or define these concepts.[44,57–58] Instead, they briefly identified actions or practices we identify as congruent with the definition of CRC we used for our analysis,[17] such as racial concordance, patient-centered communication skills, and other communication skills to address patient mistrust. These articles may be especially useful because they identify communication skills that could be developed to support CRC in practice.

## Discussion

### State of the Literature

The majority of the studies were published in 2022 with fewer studies published in 2021. This suggests an increasing research interest in CRC related to COVID-19 vaccinations as the pandemic progressed and the need for targeted interventions became more evident. A subsequent review of literature, from 2023 and forward, could provide additional insights and potentially reveal trends in CRC practices and their effectiveness over a longer period. Additionally, with many of these studies unable to measure CRC, there may be opportunities to do so given the progressive clarity in defining CRC since 2021.

Given the increasing focus on addressing disparities in healthcare, we had anticipated more articles conceptualizing CRC in this review, particularly given the absence of a clear conceptualization of the term from which specific, measurable actions can be identified, developed, and subsequently measured. Yet, this review noted few conceptual articles and commentaries, highlighting a gap in theoretical discussions and expert opinions that could help frame and guide future empirical research related to CRC and COVID-19. Of the articles reviewed, many were empirical research studies indicating a strong focus on gathering data and evidence directly from clinical or community settings. Given our focus on the COVID-19 pandemic, this makes sense.

Additionally, most of the intervention studies were public health. The one intervention study related to direct patient-PCP interaction and CRC by Lieu et al. focused on PCP outreach to patients.[50] Moreover, many of the studies adopted qualitative methods, suggesting the yet evolving science on CRC and COVID-19 screening for minoritized populations. With such a variety of research types including qualitative, quantitative, participatory action, and public health intervention studies, there seems to be a multifaceted nature to studying CRC and COVID-19 vaccinations.

Analysis also indicated an increased focus on either Black or Hispanic/Latino populations in literature published in 2022 [41, 43–45, 48–49, 51–52, 54–55] as compared to 2021 [39–40,56–59]. These specialized studies are crucial given the disproportionate impact of COVID on these minoritized groups. In 2021, there was less focus on specific minoritized populations; rather, authors addressed all racial and ethnic minoritized groups suggesting a significant push to address broader issues of racism and CRC in healthcare at the beginning of the pandemic.

None of the included articles mentioned or recommended CRC for sexual or gender minoritized groups. This gap is startling considering that sexual and gender minoritized groups suffered from more mental health disparities than their non-minoritized counterparts due to the COVID-19 pandemic, indicating the need for CRC to be studied in relation to these specific groups, in general, and in the context of COVID vaccinations.[5–10, 26–29]

## Consensus of a CRC Definition

Although difficult to ascertain, consensus on a CRC definition is important for CRC to be measured accurately (for research), to provide guidance toward greater competence for clinicians (for practice), and to provide guidance on training and development (for future practice).

Understanding the scope of how CRC is defined, allows efforts in research, practice, and future practice to be targeted to improve patient care.

Many different terms related to CRC were used in the literature reviewed. These terms aimed at describing PCPs or interventions as *culturally tailored*, *competent*, *appropriate*, *humble*, *intelligent*, *concordant*, *inclusive*, *pointed*, *grounded* and *relevant*. These terms have been described in this scoping review as similar, but in reality, they represent more than the interpersonal nature of PCP and patient interactions. Instead of interpersonal communication skills and abilities that come from intrapersonal attitudes and beliefs, terms like cultural concordance and inclusivity refer to systemic and institutional influences of health care, according to the SEM.[38] To make a clinical setting culturally concordant or inclusive may necessitate the changing of hiring practices to employ PCPs who better represent patient populations, to achieve racial concordance, or expert language interpreters, to achieve language concordance. These terms refer to potential systemic level changes, while being culturally competent, intelligent, humble, etc. refer to the attitudes and beliefs that PCPs can aim to change to influence their communication with patients at the interpersonal level. PCPs can reasonably improve or gain these skills and attitudes through training and practice to enhance their care for minoritized patient groups. With the varying terms utilized in this literature referring to different levels of the SEM, PCPs may get confused on what to prioritize in order to improve their care for all groups. A single definition with a framework of recommendations for, specifically, interpersonal patient-PCP communication can better guide PCPs to utilize CRC themselves.

A few articles used organized and informative definitions to guide PCPs on interpersonal communication skills more so than other articles. These articles were authored by Richard-Eaglin & McFarland and Singer et al. who defined cultural intelligence and culturally safe care, respectively.[47, 52] They rigorously defined these terms through a framework of capabilities or tenets throughout their respective articles. For example, Richard-Eaglin and McFarland focused on “4 capabilities of cultural intelligence”:

“Mindfulness and acceptance of differences and unique lived experiences (ie, un-conventional health-care practices, beliefs, attitudes about health and healthcare); Development of customized patient visits and treatment plans; Effective and trusting patient (person), family, and community partnerships; and Greater influence on informed patient (person) decision-making”^[47(p8)]^

Singer et al. divided their definition of culturally safe care in 5 clear-cut tenets: 1. Partnerships; 2. Personal activities of daily living; 3. Patient centering; 4. Prevention of harm; and 5. Purposeful self-reflection.[52] Even a study that never explicitly mentioned a CRC term, by Opel et al., was able to provide a generalizable list of important communication skills that CRC partly addresses: 1. Lead with listening; 2. Tailor responses to patient concerns; 3. Briefly describe the regulatory and developmental process surrounding COVID-19 vaccines using accessible language; and 4. Acknowledge uncertainty.[57] These articles incorporate frameworks in a manner that is clear and utilizable for research, PCP utilization, and training regarding COVID-19 vaccinations.

While several articles clearly defined CRC, some misdefined CRC-related terms or failed to provide a definition, potentially causing problems in definition consistency for research, training, and evaluation.[39–40, 43–44, 48, 57–59] Further, multiple authors limited their definitions of cultural competency to reflect language and racial concordance only.[42, 46, 49, 53, 58] For example, Leibowitz et al. defined cultural competency as “leveraging established patient-provider relationships, using interpreters, and involving staff who reflected the diversity of the communities”. ^[42(p1)]^ Here, authors refer to language and racial concordance which reflect changes that must be made to the systemic and institutional levels of the SEM, not changes that PCPs can make with their communication via cultural competency. Cultural competency refers to a wider range of PCP skills and attributes that involve the humility and understanding of other cultures.[17] Several articles failed to provide any definition of CRC, only providing recommendations of actions aligned to CRC.[39–40, 43–44, 48, 54–55, 58–59] This inconsistency of definitions, referring to different SEM levels, is likely to confuse researchers and PCPs who want to study and utilize CRC.[17] With such a lack of concept clarity within the literature, it is important that authors, researchers, and PCPs are diligent in defining what it means to utilize CRC-related concepts.

Of note, half of the articles that did define CRC-related terms were based on community perspectives and often mentioned language or racial concordance exclusively.[48–51, 53, 60] Some of these articles failed to use the word “concordance” but included enough content for the study reviewers to assume its relevance to the term. The studies that focused on capturing a specific community’s perspective may also offer valuable insights for CRC utilization in specific populations. For instance, Castellon-Lopez et al. defined their very own “culturally-pointed” strategy based on survey and interview findings from a predominantly Latino community.^[49(p8)]^ They suggested that trusted PCPs should focus on providing evidence-based recommendations and dispelling myths of the COVID vaccination rather than trying to add to the “rapid fire culturally targeted content” that Latino communities may not trust.^[49(p8)]^ This unique understanding gives light to the importance of gathering community insights. The incorporation of feedback from minoritized communities in the design and implementation of CRC studies and interventions is clearly crucial for their success in specific communities.[17, 27] However, this may lead to a dilemma of having specific CRC definitions by community or having a single agreed upon definition for all communities. CRC frameworks may be able to address this dilemma by mentioning the importance and possible methodologies of gathering community perspectives through the integration of other frameworks (e.g. participatory action action research, community-based participatory research, or community-oriented primary care).

In the literature reviewed, applied terminology such as cultural *competence*, *humility*, and *concordance* could be interpreted in different ways and apply to different levels of the SEM. Some of the articles used these terms interchangeably or in overlapping ways, leading to obvious confusion and inconsistencies. There is no one commonly applied term or definition across articles in the context of COVID-19 vaccination. These different conceptualizations make it hard to measure (research), hard to provide guidance toward greater competence for clinicians (practice), and hard to provide guidance on training and development (future practice). It is critical that we establish a standardized definition and framework for CRC to ensure its consistent application in research, practice, and future practice while also attending to the needs of specific communities.[17]

## Measuring CRC in Clinical Settings

Many of these articles took a population-based or community-oriented approach when referring to CRC-related terms rather than focusing solely on direct patient-provider communication, indicating a gap in research specific to clinical settings.[39–40, 42, 44–47, 49, 52, 56, 57–58] A study that most closely represented a test of the effects of PCPs utilizing face-to-face CRC was by Leibowitz et al.[42] As mentioned previously, this study team used health centers that fostered trust and cultural competence by utilizing established PCP relationships, interpreters, and racial concordance. However, their only assessment for cultural competence was through an unknown cultural competence survey answered by half their patient participants. With the absence of other statistics, details, or patient insights, their only CRC-related evaluation was that “providers felt that trust and cultural competency were key to the initiative’s success”.^[42(p3)]^ Future research focused on CRC would require more nuance and advanced methods, specifically quantitative measures of CRC implementation in clinical setting and corresponding outcomes.

An example of a more measurable method of researching CRC can be found in a randomized controlled trial by Lieu et al.[50] The study team sent a culturally tailored, COVID-19 vaccine outreach messages from PCPs to their patients and compared these patients to two other patient groups: patients receiving standard clinical care messages and patients receiving no provider messages. The culturally tailored message addressed potential cost and immigration status issues as well as racial and ethnic COVID-19 disparities in writing. The messages were further tailored—including photos of older adults of the respective patients’ racial and ethnic backgrounds. With this clear difference of exposure between the three patient groups, Lieu et al. was able to directly assess and compare COVID-19 vaccination rates. They found that the difference between the groups receiving messaging was not significant, but that both messaging groups had significantly higher vaccination rates than the no messaging group. Although this study does not implicate the importance of CRC, it provides a methodology that accurately tests outcomes of CRC in practice.

## Training CRC for PCPs

Prior to evaluating CRC efficacy and effectiveness in clinical settings, focus must also be placed on training PCPs to implement CRC principles. This training should center on authentic community perspectives and real-world clinical scenarios to equip providers for culturally responsive care. This involves inviting community advocates to share concerns, values, and health-related experiences that shape how their communities engage with healthcare. Including voices from various social identities offers a nuanced view of the challenges and barriers many patients face. Effective training should feature perspectives from PCPs, with example clinical scenarios that help participants empathize with patients’ experiences and understand how cultural dynamics affect clinical interactions. These scenarios can allow providers to practice applying CRC in realistic contexts, making the training both practical and directly relevant to everyday healthcare.

To further enhance the training, it should include contributions from experts in CRC and related concepts. These experts can present a range of strategies for understanding and implementing CRC along with insights into the ethical and social dimensions of healthcare.

Ideally, the training would also incorporate diverse learning formats, such as interactive presentations, CRC utilization in published articles, and relevant supplemental resources to deepen understanding and foster skill-building. Finally, a strong CRC training program should be ongoing rather than a one-time event. Follow-up sessions, self-assessment tools, and access to additional resources can help providers continually reflect on and improve their CRC skills, ultimately leading to more equitable and compassionate patient care. This guidance has been primarily informed and utilized by the recent construction of our own CRC medical education training series led and described by Ward et al.[28–30]

## Limitations

The majority of articles were published in 2022, which indicates a potential publication year bias towards more recent research. However, this may have been unavoidable due to the recent emergence of the COVID-19 pandemic and subsequent vaccine rollout. This limits our understanding of older CRC-related studies that may have defined and recommended it more clearly. With such a limited timeframe, we were also unable to capture the long-term trends or the full evolution of CRC practices.

Even though we tried to focus on direct patient-PCP CRC, most studies only briefly incorporated direct communication because they were primarily based on public health messaging and advocacy. This may have been due to the inability or fear patients had when asked to physically visit clinical settings during the height of the COVID-19 pandemic. This literature focus may have diluted the specific insights relevant to in-person clinical interactions, but it also highlights the need for more research on the implementation and measurement of CRC within clinical interactions. Ongoing work by Ward et al., which proceeds the dates of literature reviewed, is advancing the conceptualization of CRC and should be considered in future reviews.[17, 26–33] This work challenges existing definitions and conceptualizations of CRC.

## Conclusion

This scoping review provides a comprehensive examination of CRC between patients and PCPs in the context of COVID-19 vaccination and booster screening for racially, ethnically, sexually, and gender minoritized patients. The literature reveals a recent increase in research interest, particularly empirical studies, highlighting the need for more conceptual and theoretical work to standardize CRC definitions and practices. Many studies published in 2022 focus on Black and Hispanic/Latino populations, underscoring the disproportionate impact of COVID-19 on these groups, while also highlighting a need for more research on sexual and gender minoritized groups who face significant health disparities.

Applied terminology related to CRC varied widely, inviting potential confusion and underscoring the need for a single, comprehensive definition and framework. Establishing a standardized CRC framework is essential for improving health outcomes, guiding clinical practice, and ensuring equitable care for all minoritized communities. Additionally, incorporating community insights is crucial for the success of CRC interventions. Our review provides a snapshot, in-time, of the application of CRC in the context of COVID-19. This snapshot highlights the need for continued advancement in conceptualizing CRC in order to develop future research methods, clinician training, and practice guidance.

## Data Availability

All relevant data are within the manuscript.

